# Spread and control of COVID-19 in China and their associations with population movement, public health emergency measures, and medical resources

**DOI:** 10.1101/2020.02.24.20027623

**Authors:** Songmin Ying, Fei Li, Xinwei Geng, Zhouyang Li, Xufei Du, Haixia Chen, Sisi Chen, Min Zhang, Zhehua Shao, Yinfang Wu, Madiha Zahra Syeda, Fugui Yan, Luanqing Che, Bin Zhang, Jian Lou, Shaobin Wang, Zhengming Chen, Wen Li, Ye Shen, Zhihua Chen, Huahao Shen

## Abstract

**BACKGROUND:** The COVID-19 epidemic, first emerged in Wuhan during December 2019, has spread globally. While the mass population movement for Chinese New Year has significantly influenced spreading the disease, little direct evidence exists about the relevance to epidemic and its control of population movement from Wuhan, local emergency response, and medical resources in China.

**METHODS:** Spearman’s correlation analysis was performed between official data of confirmed COVID-19 cases from Jan 20^th^ to Feb 19^th^, 2020 and real-time travel data and health resources data.

**RESULTS:** There were 74,675 confirmed COVID-19 cases in China by Feb 19^th^, 2020. The overall fatality rate was 2.84%, much higher in Hubei than in other regions (3.27% vs 0.73%). The index of population inflow from Hubei was positively correlated with total (Provincial r=0.9159, p<0.001; City r=0.6311, p<0.001) and primary cases (Provincial r=0.8702, p<0.001; City r=0.6358, p<0.001). The local health emergency measures (eg, city lockdown and traffic control) were associated with reduced infections nationwide. Moreover, the number of public health employees per capita was inversely correlated with total cases (r=−0.6295, p <0.001) and infection rates (r =−0.4912, p <0.01). Similarly, cities with less medical resources had higher fatality (r =−0.4791, p<0.01) and lower cure rates (r = 0.5286, p<0.01) among the confirmed cases.

**CONCLUSIONS:** The spread of the COVID-19 in China in its early phase was attributed primarily to population movement from Hubei, and effective governmental health emergency measures and adequate medical resources played important roles in subsequent control of epidemic and improved prognosis of affected individuals.

## INTRODUCTION

In mid-December 2019, an unexplained mass of pneumonia cases occurred in Wuhan, Hubei province of China^1^. Early epidemiological investigations indicated that the cause of the infection could be linked to the Wuhan South China Seafood Market^2^. High-throughput sequencing revealed a novel beta-coronavirus that was provisionally called 2019 novel coronavirus (2019-nCoV)^3,4^, which has now been officially renamed to COVID-19 by WHO^5,6^. A number of studies showed that the epidemiological, clinical, laboratory, and radiological features of COVID-19 are similar, albeit less deadly, to those of severe acute respiratory syndrome coronavirus (SARS) in 2003 and Middle East respiratory syndrome (MERS) in 2012^7-9^, and evidences pointing to the human-to-human transmission in hospital and family settings have now been firmly established^10^.

Due to the Chinese Lunar New Year travel rush, the COVID-19 epidemic has gradually spread across the country and even worldwide within a limited time frame^11^. In response to the situation, unprecedented measures have been taken by central and local government to contain the outbreak and prevent its further spread across China. On Jan 23^th^, the Wuhan City Epidemic Prevention and Control Headquarters announced that all urban buses, subways, ferries, and long-distance passenger operations were suspended, and that the passages of airports and train stations were temporarily closed^12^. Subsequently, major cities within Hubei province started to implement lockdown on Jan 26^th^ or 27^th^, 2020, except the remote Shennongjia Forestry District due to the very limited number of COVID-19 cases. Unfortunately, around five million people had already left Wuhan by then since the emergence of COVID-19^13,14^. As the situation continued to deteriorate throughout China, the WHO declared it as a global public health emergency on Jan 30^th^ 2020^15^.

Several studies have already reported on the molecular, clinical and epidemiological features of COVID-19^11,16-18^. However, to date no study has quantified the role of population movement in the spread of epidemics across different parts of China, or assessed the effectiveness of local public health emergency response, and medical resources on control of epidemics and prognosis of the patients. To help fill the evidence gap, we presented detailed analysis of available data of reported cases from Jan 20^th^ to Feb 19^th^, 2020 in China, along with information related to population travel, public health emergency measures, and available medical resource from various regions of China. The main objectives of this study were to present a real-world paradigm of the importance of governmental health emergency strategies in subsequent control of epidemic and the local medical resources in association with the prognosis of affected cases.

## METHODS

### COLLECTION OF EPITHELIAL DATA

The daily data of confirmed COVID-19 cases in various regions of China from Jan 10^th^ to Feb 19^th^, 2020, were obtained from National or Provincial Health Commission in China (NHC/PHC). Data of global COVID-19 cases were collected from WHO. It included the daily new and cumulative cases of confirmed patients, cured patients, and deaths. All cases included detailed epidemiological history and the dates at which incidents occurred. Provinces with small number of cases or heavily weighted with incomplete exposure history cases, such as Jiangxi province, were excluded.

### COLLECTION OF MEDICAL RESOURCES DATA

Information on medical resources were extracted from the national and local Statistical Yearbooks in 2019, which included data on number of hospitals, health workers, and hospital beds per 1,000 population, health expenditure per capita, and local population size. After excluding those with incomplete data in the Statistical Yearbooks, 9 cities of Hubei Province and 20 cities from other 14 provinces of China were finally included in this study (Table S1).

### POPULATION OUTFLOW/INFLOW INDEX

Data on population movement were retrieved from the Chinese Lunar New Year Travel Information, which was released daily by Baidu Migration Map (http://qianxi.baidu.com). We obtained the daily outflow index in Hubei that occurred from Jan 1^st^ to Feb 19^th^, 2020, which were matched with same data in the previous year according to lunar dates for a direct comparison. Also, we obtained the proportion of the daily outflow index from Hubei to other provincial areas and 51 cities which provided detailed exposure history for the confirmed cases from Jan 10^th^ (the start of the Lunar New Year travel rush) to Jan 26^th^, 2020 (lockdown of major cities in Hubei).

### DAILY GROWTH RATE OF SECONDARY CASES

Confirmed cases were categorized into two groups, the primary (with clear history of staying or traveling in Hubei province within one incubation period) and secondary (those not known to be primary) cases, by two independent researchers in a blinded manner. The daily growth rate of secondary cases was calculated as new secondary daily cases divided by the cumulative number of the day before. The lag time between primary and secondary cases was identified by using the displacement with the highest correlation from the cross-correlation result.

### STATISTICAL ANALYSIS

The daily inflow index of certain provinces and cities was calculated by multiplying the daily outflow index within Hubei and the corresponding proportion. We defined the total inflow index as the sum of daily inflow index from Jan 10^th^ to 26^th^, 2020. We used principal components analysis to reduce the dimensionality of five initial parameters of medical resources, and to further obtain the synthetic score of these parameters^19^. Factor loadings for concordance and overall satisfaction were low and thus were removed. Table 2 shows 4 item factor loadings for the final two-factor solution, which explained 96% of the variance. Medical resource scores equal to comp1*proportion1 plus comp2*proportion2 (Table S2).

The correlations between the number of total confirmed or primary cases and total inflow index at provincial or city scale, between the medical resources score and fatality or cure rates, and between the employees in centers for disease control and prevention (CDC) per capita and the confirmed cases or the incidence of COVID-19 were analyzed using Spearman’s correlation analysis. Cross-correlation of primary and secondary cases was calculated by Pearson’s correlation analysis. Principal components analysis was performed on Stata 14.0, and other data were analyzed using Prism GraphPad 8.0. Statistical significance was set at *p* < 0.05.

## Results

### Time trend of COVID-19 epidemics in China

As of Feb 19^th^, 2020, a total of 74,675 confirmed COVID-19 cases had been reported in China, with 83.1% (62,031) being in Hubei province (Fig. 1A). The cumulative number of confirmed cases was below 1,000 before Jan 23^rd^, 2020, and increased by almost ten-fold by Jan 30^th^, 2020. There was a further three-fold increase in the number of confirmed cases by Feb 6^th^, 2020, which continues to grow until now, but at much slow pace. In Hubei province the number of daily confirmed cases reached peak on Feb 4^th^, 2020 (Fig. 1B), while in other regions, it reached a plateau on Jan 30^th^, 2020, and started to decline from Feb 3^rd^, 2020 (Fig. 1C).

**Figure 1.**
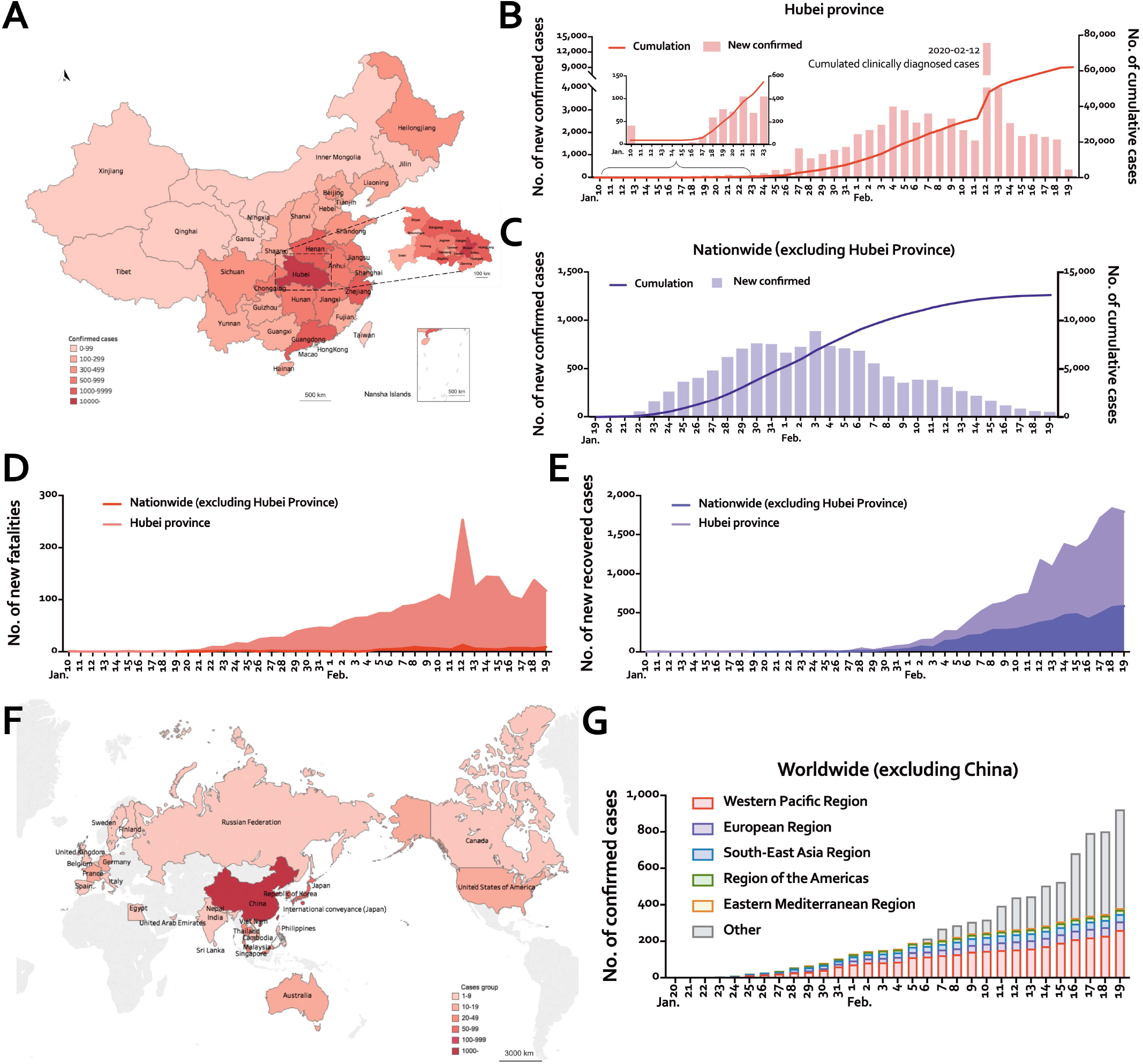
Epidemiological Features of COVID-19 in Hubei, China, and Worldwide. (A) The spatial distribution of 74675 cases with confirmed COVID-19 infection on Feb19^th^, 2020 in China. The color of regions represents the number of confirmed cases. Magnified image shows the spatial distribution of 62031 confirmed cases in cities and regions of Hubei province. (B-C) Time course of the newly confirmed COVID-19 cases in Hubei province (B) and in China excluding Hubei (C). (D-E) Daily number of death (D) and cure (E) of COVID-19 patients in Hubei province and in China excluding Hubei. (F) Global distribution of countries, territories, or areas with confirmed COVID-19 patients Feb19^th^, 2020. (G) Time course of cumulative confirmed COVID-19 cases (n=924) worldwide excluding China.

As of Feb 19^th^, 2020, the overall case fatality rate was 2.84%, much higher in Hubei province than in other regions (3.27% vs 0.73%), with Hubei accounted for 95.7% (2029/2121) of total deaths nationwide. There was irregularity in the reported daily number of deaths over time, with a sudden rise on Feb 12^th^, 2020, coinciding with change of diagnostic criteria (Fig. 1D). The daily number of cured cases has continuously increased both in Hubei province and in other parts of China (Fig. 1E). Following the first confirmed COVID-19 case outside of China in Thailand, as of Feb 19^th^, 2020, a total of 924 cases have been reported worldwide excluding China (Fig. 1F). However, the daily cumulative confirmed cases worldwide excluding China grew slowly (Fig. 1G).

### Correlation of population movement with the COVID-19 epidemic

In both 2019 and 2020, the daily population outflow from Hubei started to rise steadily for 7-10 days before the Lunar New Year’s Day (Jan 25^th^, 2020) (Fig. 2A). However, starting on Jan 20^th^, 2020, there was a sudden surge in the outflow index when it was acknowledged publicly by the government the fact of human-to-human transmission. In contrast, there has been a dramatic decrease in the outflow index since Jan 26^th^, 2020, compared with that of 2019, following the total lockdown of most cities, first in Wuhan and then elsewhere, in Hubei province (Fig. 2A).

**Figure 2.**
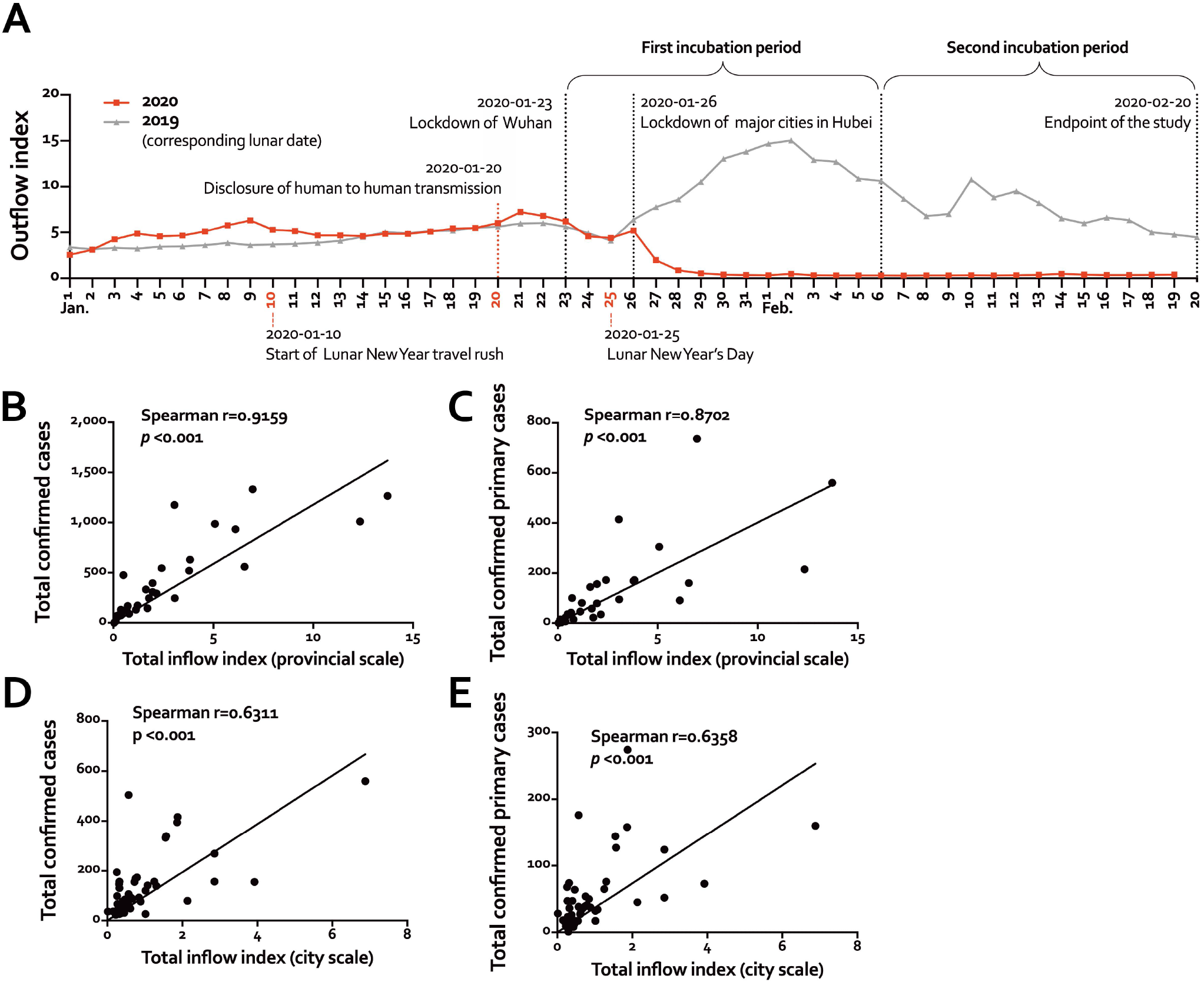
Associations of Population Movement with the COVID-19 Prevalence in Other Chinese Areas. (**A**) Outflow index of Hubei province during period of Jan 1^st^ to Feb 19^th^ in 2019 and 2020; (B-E) The correlations between the total number of or primary confirmed COVID-19 cases and the total index of inflow at the provincial (B, C) and city (D, E) scale.

We correlated the daily inflow index of 30 provincial areas and 51 cities in 18 of these provinces with the number of reported total or primary cases in the same areas and found a very strong correlation both at province and city levels. The index of population inflow from Hubei province strongly positively correlated with number of confirmed total (Provincial scale: r=0.9159, p<0.001, Fig. 2B; City scale: r= 0.6311, p<0.001, Figure 2D) and primary cases (Provincial r= 0.8702, p<0.001, Fig. 2C; City r= 0.6358, p<0.001, Fig. 2E).

### Growth rate of secondary case across different regions and cities

Overall, the ratio of secondary to primary cases (S/P ratio), a simple index for measuring the growth of an epidemic, varied greatly across different provincial areas (Fig. 3A), with Heilongjiang (10.7) and Hong Kong (5.5) being the highest, and Tibet (0) and Qinghai (0.2) being the lowest. However, there was little correlation between total number of confirmed cases and S/P ratio. For example, Guangdong had the largest number of confirmed cases (1,332) but very low S/P ratio (0.35).

**Figure 3.**
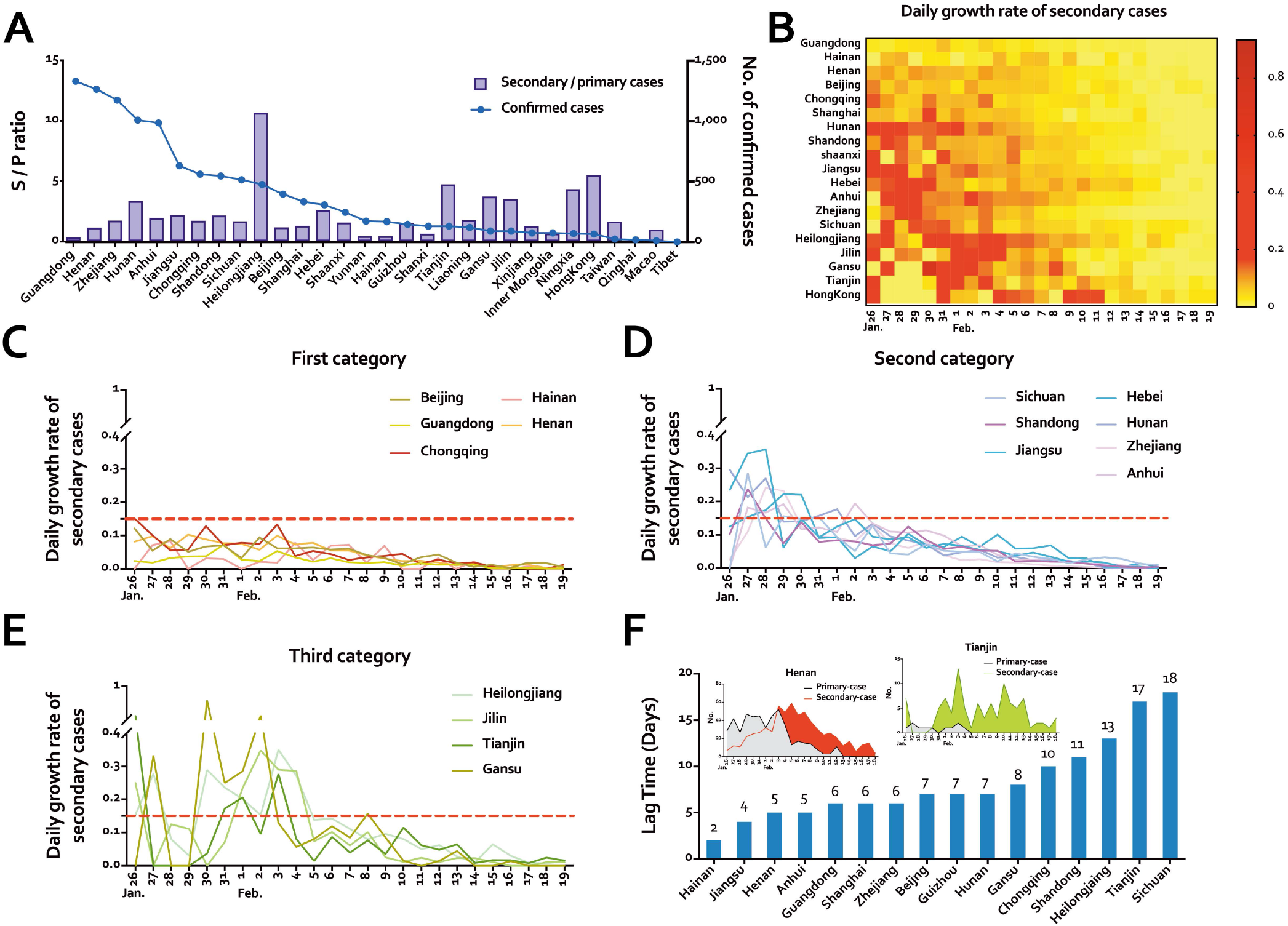
Secondary Case Growth Rate and Lag Time in other Chinese Regions. (A) Cumulative number of confirmed cases and ratio of secondary to primary cases (S/P ratio) in provinces/cities/region by Feb 19^th^, 2020. (B) Daily growth rate of secondary cases in provinces/cities/regions from Jan 26^th^ to Feb 19^th^, 2020. (C-E) Three types of provincial administrative areas depending on variety of daily growth rate of secondary cases. (F) Lag time between primary and secondary cases in various provinces and examples of Jiangsu (∼ 4 days) and Tianjin (∼ 17 days) from Jan 26^th^ to Feb 19^th^, 2020.

Fig. 3B shows the heat map of the daily growth rate of secondary cases from Jan 26^th^ to Feb 19^th^, 2020 in different provinces. We found although daily growth rate varied among different provincial areas, some of them had common characteristics (Fig. 3B). Based on the daily growth rate of secondary cases we further divided study regions into three categories. In the first group the maximum values for daily growth rates were all <1.5, including Beijing, Guangdong, and Chongqing (Fig. 3C). Second category of provinces/cities (eg, Sichuan, Hebei, and Zhejiang) showed much higher growth rate in the first few days, from Jan 26^th^ to 30^th^, 2020, followed by a downward trend until reaching that of the first category (Fig. 3D). In the last category (eg, Heilongjiang and Tianjin) the daily growth rate showed a sustained and rapid rise, especially during the 5 days after Jan 30^th^, 2020 (Fig. 3E).

We further examined the lag time, or displacement, between primary and secondary cases by area, which could reflect the time delay in implementing effective local containment measures. Although the lag time varied across different areas, most appeared to be about 1 week (Fig. 3F), with exceptions such as Jiangsu, Henan, Tianjin, and Heilongjiang. In Jiangsu province (631 confirmed cases) and Henan province (1265 confirmed cases), the lag time was only about 4 and 5 days, respectively; while in Heilongjiang (476 confirmed cases) and Tianjin (130 confirmed cases) it was approximately 13 and 17 days, respectively.

### Correlation of prognosis and transmissibility of COVID-19 with medical resources

Table S3 shows the summarized data of the numbers of CDC employees and severity of local epidemic. Overall the provinces with higher number of CDC employees per 1,000 population tended to have fewer confirmed cases (r = −0.6295, p < 0.001, Fig. 4A) and lower infection rate (r =−0.4912, p < 0.01, Fig. 4B). Moreover, in the principal components analysis of the correlation between the capacity of medical resources and the trends of fatality and cure rates, we found that the cities with limited medical resources tended to have higher case fatality (r = −0.4791, p < 0.01, Fig. 4C) and lower cure rates (r = 0.5286, p < 0.01, Fig. 4D).

**Figure 4.**
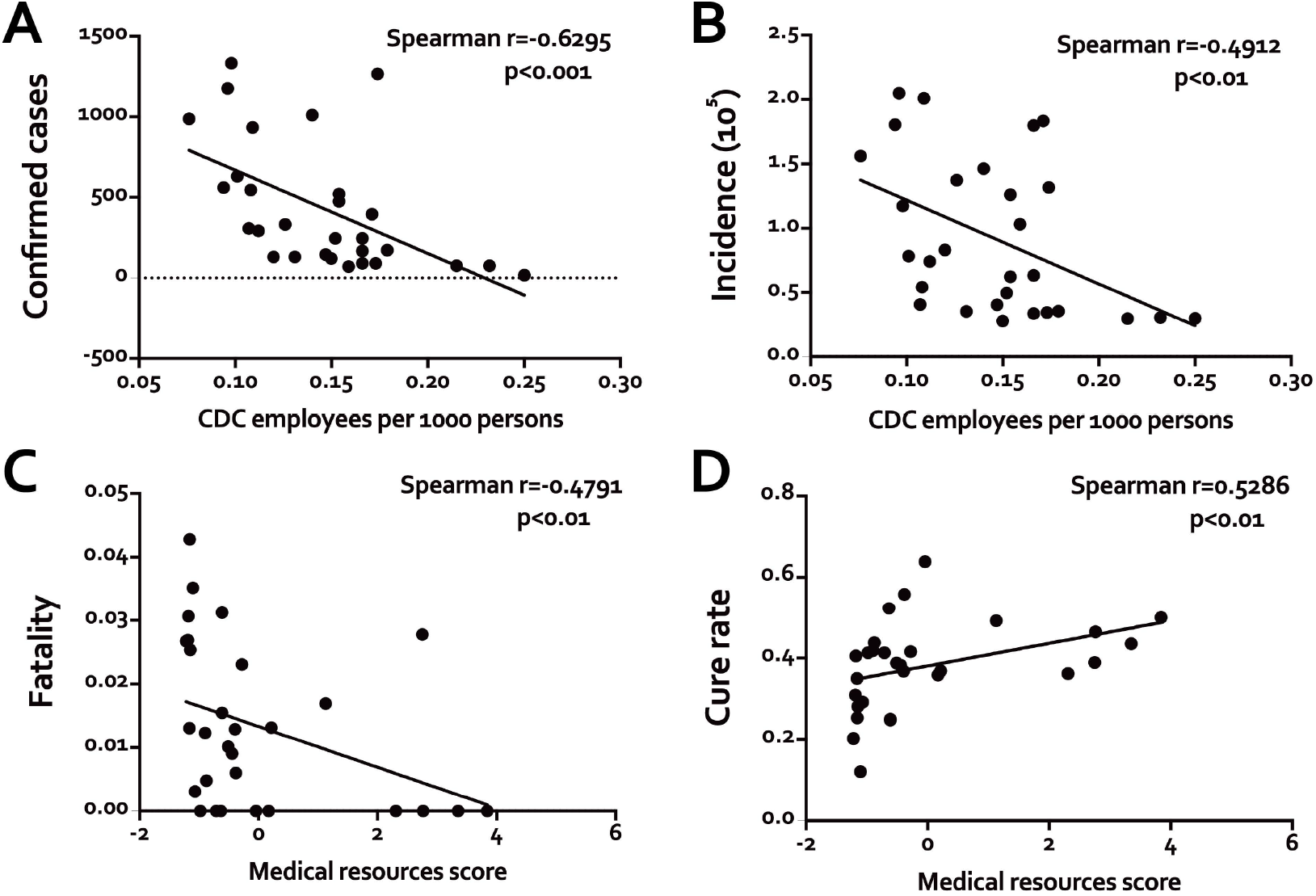
Correlations between the Abundance of Medical Resources and Disease Prognosis. (A, B) The correlations between confirmed cases (A) or incidence (B) and CDC employees per 1000 persons of provinces in China (excluding Hubei and Tibet). (C, D) Correlations between the medical resources scores and the fatality (C) or the cure rate (D) of certain Chinese cities.

## DISCUSSION

This study presented detailed analyses of time trends of COVID-19 epidemic across different parts of China and their associations with population movement, public health emergency measures, and medical resources. We showed that the rapid spread of the COVID-19 epidemic across China was strongly associated with the mass population movement out of Hubei province, particularly Wuhan city, before the Chinese Lunar New year, which was subsequently disrupted effectively by the total lockdown of Wuhan and other cities in Hubei provinces. Although there were variations in the pace of control across different regions of China, the epidemic outside of Hubei province was contained rapidly and effectively through various public health emergency measures. As well as public health measures, local capacity including the number of public health staff and available medical resources also played important roles in control of epidemic and improved prognosis of affected individuals.

The first case of novel COVID-19 infection was reported during early December 2019 and it was not publicly acknowledged until 20 January 2020 that the virus could be transmitted from human to human, which triggered rapid and drastic public health measures both in Hubei and rest of China to try to contain the spread of virus. The total lockdown of Wuhan city on 23^rd^ January 2020, followed by other cities in Hubei a few days later, appeared too late to prevent the epidemic from spreading into other regions of China, for by then over 5 million people had already left Wuhan. However, without such drastic measures, the situation could be much worse. The data from Baidu Migration Map showed that mass population movements out of Wuhan and Hubei province took place not only before but throughout the whole of Chinese Lunar New Year period. Based on comparison with 2019 data, without lockdown an additional 15 million people could have traveled from Wuhan to other regions (and overseas) plus similar or even larger number who would have travelled from other regions to Wuhan. Moreover, there would have been massive internal population movement within Wuhan and other cities in Hubei during the same period, further exacerbating the epidemic.

Expectedly the severity of COVID-19 epidemic outside of Hubei province, especially during the initial phase, was strongly related to the scale of inward population movements from Wuhan. However, the epidemic was rapidly brought under control in most areas by introduction of various public health emergency measures, as demonstrated by decrease of the daily number of confirmed cases starting from the 1^st^ or 2^nd^ week of February and the lack of clear correlation between S/P ratio and number of total confirmed cases. Despite this, the pace with which the epidemic was contained varied across different areas as assessed by various parameters examined, including S/P ratio, the daily growth rate of secondary cases and lag time. For example, in Guangdong and Beijing, both of which were badly affected by SARS outbreak in 2003, the epidemic was effectively contained at very early stage, suggesting adequate level of local preparedness and experience in dealing with such epidemics. Similarly, in Zhejiang province, doctors can pre-screen suspected patient through the internet application, which greatly reduce the probability of hospital transmission. Moreover, using cloud computing facilities and integrated big data platform, public health doctors in Zhejiang province was also able to cross-examine every suspected case. These measures have contributed importantly to a sharp and continual downward trend in the number infected after Jan 30^th^, 2020. On the other hand, in several other areas (eg, Heilongjiang and Tianjin) there were prolonged delays in containing the epidemic, reflecting probably less effective local measures in controlling the epidemic. For example, in Heilongjiang, the S/P ratio approached 11, which was the highest across all regions outside of Hubei, suggesting nearly 90 percent of the confirmed cases resulted from family gatherings.

The regional variations in the pace of epidemic containment was also evident by comparison of the mean lag time between primary and secondary cases. Overall the mean lag time was about 1 week, with particularly low value in Jiangsu (∼ 4 days) and Henan (∼5 days), and particularly high value in Heilongjiang (∼13 days) and Tianjin (∼17 days). Henan is a neighboring province of Hubei with a total population of 100 million people and extensive transport connections with Wuhan. In recognition of forthcoming epidemic, the Henan provincial government introduced strong measures to greatly reduce and restrict public transport from Wuhan areas into Henan even before the total lockdown of the Wuhan city.

Although the COVID-19 shared many similar epidemiological features to those of SARS in 2003^16^, it appeared to be much less deadly^20,21^, with the overall case fatality rate of less than 3% as opposed to ∼10% for SARS. However, there was great difference in the case fatality rates between Hubei, particularly Wuhan and rest of the China. As the epicenter of COVID-19 outbreak, medical and health services in Hubei Province were overwhelmed and ill prepared for such a rapid and substantial increase in the number of infected cases, leading to poor and inadequate management of patients, hence poor prognosis. Apart from difference in the capacity of medical services, other factors, including age of people affected and proportion with other comorbidities may also contribute to the higher case fatality observed in Wuhan and Hubei. However, without detailed clinical data from individual patients, we were not able to examine these issues directly. It is also possible that a large number of minor cases were not promptly detected or diagnosed in Wuhan and Hubei, resulting in higher case-fatality rate. Indeed, as the medical service started to improve gradually and large number of cases were properly diagnosed the case fatality rates had started to decreased steadily over the last two weeks. Outside of Hubei, although the case fatality rates were very low, we also found a significant correlation between health scores and overall prognosis of patients. In recognizing the burden of epidemic in Hubei province and need for providing prompt and adequate medical service to those infected, the Chinese government has created a “province to city” support system, in which each city in Hubei province received direct and targeted support from at least one appointed province.

In summary the present study showed that the spread of the COVID-19 epidemic in China (and elsewhere in other countries) could be attributed primarily to the mass population movement from Hubei prior to the Chinese Lunar New Year. Subsequently, effective governmental health emergency measures introduced both in Hubei and elsewhere have played important roles in rapid and effective control of the epidemic in China. Although many other unmeasured factors, such as local climates and characteristic of individuals affected, may explain part of our findings, the present study also provided good evidence that adequate levels of investments in public health (eg, number of public health staff) and medical resources can lead to improved control of epidemic and better prognosis of the infected individuals. Despite the rapid improvement, the COVID-19 epidemic in China and elsewhere is not yet over and vigorous public health measures are still warranted in order to totally eliminate the epidemic.

## Data Availability

No external datasets or supplementary material

## FOOTNOTES

Partially supported by the Zhejiang University Special Scientific Research Fund for COVID-19 Prevention and Control.

## APPENDIX

The authors’ affiliations are as follows: the Key Laboratory of Respiratory Disease of Zhejiang Province, Department of Respiratory and Critical Care Medicine, Second Affiliated Hospital of Zhejiang University School of Medicine (S.Y., F.L., X.G., Z.L., X.D., H.C., S.C., M.Z., Z.S., Y.W., M.Z.S., F.Y., L.C., B.Z., J.L., S.W., W.L., Z.C., H.S.), and the Hangzhou Mitigenomics Technology Inc. (YS) – all in China, and the Clinical Trial Service Unit & Epidemiological Studies Unit, Nuffield Department of Population Health, University of Oxford, United Kingdom (CZ).

---

**Table S1.** Indexes of Health Resources for Included Cities

| City | Hospitals per<br>1000 persons | Health<br>workers per<br>1000 persons | Beds per<br>1000 persons | Health<br>expenditure<br>per capita | Population<br>(1000 persons) | Confirmed<br>cases | Fatality | Cure rate |
| --- | --- | --- | --- | --- | --- | --- | --- | --- |
| *Wuhan | 0.036 | 12.30 | 8.60 | 3522 | 11081 | 45027 | 3.52% | 12.10% |
| *Xiaogan | 0.012 | 4.92 | 4.63 | 2066 | 4920 | 3329 | 2.67% | 20.22% |
| *Huanggang | 0.011 | 5.30 | 5.68 | 2403 | 6330 | 2839 | 3.06% | 40.47% |
| *Huangshi | 0.015 | 7.72 | 6.36 | 2890 | 2470.7 | 967 | 2.69% | 30.92% |
| *Jingmen | 0.021 | 7.41 | 5.82 | 3244 | 2896.5 | 794 | 4.28% | 25.31% |
| *Xianning | 0.013 | 8.68 | 5.76 | 1298 | 2543.3 | 766 | 1.31% | 34.99% |
| *Tianmen | 0.012 | 6.47 | 4.76 | 515 | 1273.5 | 473 | 2.54% | 28.12% |
| *Shiyan | 0.018 | 8.31 | 8.73 | 1974 | 3406 | 641 | 0.31% | 29.17% |
| *Enshi | 0.011 | 5.44 | 5.97 | 978 | 4020.4 | 244 | 1.23% | 41.80% |
| Wenzhou | 0.016 | 7.21 | 4.59 | 2711 | 9250 | 504 | 0.00% | 41.27% |
| Shenzhen | 0.011 | 8.82 | 3.65 | 1465.45 | 13020 | 416 | 0.48% | 43.75% |
| Chongqing | 0.026 | 8.79 | 7.10 | 3836.11 | 31017.9 | 552 | 0.91% | 38.22% |
| Shanghai | 0.015 | 8.52 | 10.10 | 8630.3 | 24240 | 333 | 0.60% | 55.86% |
| Beijing | 0.034 | 16.33 | 5.73 | 10106.42 | 21542 | 395 | 1.01% | 38.73% |
| Fuyang | 0.015 | 5.04 | 4.10 | 1688.7 | 8207 | 155 | 0.00% | 41.29% |
| Xinyu | 0.014 | 7.81 | 5.00 | 2094.23 | 1160.8 | 130 | 0.00% | 52.31% |
| Harbin | 0.029 | 8.15 | 8.57 | 3154.5 | 1085.8 | 194 | 1.55% | 24.74% |
| Bengbu | 0.034 | 7.27 | 5.40 | 1774 | 3392 | 160 | 3.13% | 25.00% |
| Ganzhou | 0.050 | 5.28 | 4.89 | 1551.05 | 8507.5 | 76 | 1.32% | 36.84% |
| Nanyang | 0.052 | 7.93 | 4.70 | 2174.6 | 10013.6 | 155 | 1.29% | 36.77% |
| Hangzhou | 0.032 | 14.69 | 10.49 | 2929 | 9806 | 169 | 0.00% | 63.91% |
| Xi'an | 0.035 | 14.07 | 7.77 | 2048.8 | 10003.7 | 120 | 0.00% | 35.83% |
| Qingdao | 0.045 | 8.94 | 6.82 | 794 | 9394.8 | 59 | 1.69% | 49.15% |
| Jinan | 0.033 | 13.98 | 8.76 | 1513.5 | 7460.4 | 47 | 0.00% | 36.17% |
| Shijiazhuang | 0.023 | 9.61 | 5.54 | 1122.88 | 10951.6 | 28 | 0.00% | 46.43% |
| Guiyang | 0.039 | 10.13 | 7.73 | 2706.47 | 4881.9 | 36 | 2.78% | 38.89% |
| Bijie | 0.041 | 4.86 | 5.70 | 1526.23 | 6686.1 | 23 | 0.00% | 43.48% |
| Zunyi | 0.066 | 9.67 | 7.80 | 3100 | 6270.7 | 32 | 0.00% | 50.00% |
| Tianjin | 0.037 | 6.48 | 4.38 | 5554.36 | 15568.7 | 130 | 2.31% | 41.54% |

\*Cites of Hubei Province

**Table S2.** The Results of Principal Component Analysis (PCA)

| Variable | Component 1 | Component 2 | Component 3 | Component 4 |
| --- | --- | --- | --- | --- |
| Hospitals per 1,000 persons per case | 0.485 | 0.015 | 0.757 | 0.436 |
| Health workers per 1,000 persons per case | 0.491 | 0.056 | -0.649 | 0.570 |
| Beds per 1,000 persons per case | 0.510 | 0.022 | -0.047 | -0.487 |
| Health expenditure per capita | 0.510 | 0.022 | -0.047 | -0.487 |
| Population | -0.057 | 0.998 | 0.027 | -0.018 |
| Eigenvalue | 3.820 | 0.992 | 0.167 | 0.021 |
| % Variance Proportion | 76.41 | 19.83 | 3.34 | 0.42 |

**Table S3.**
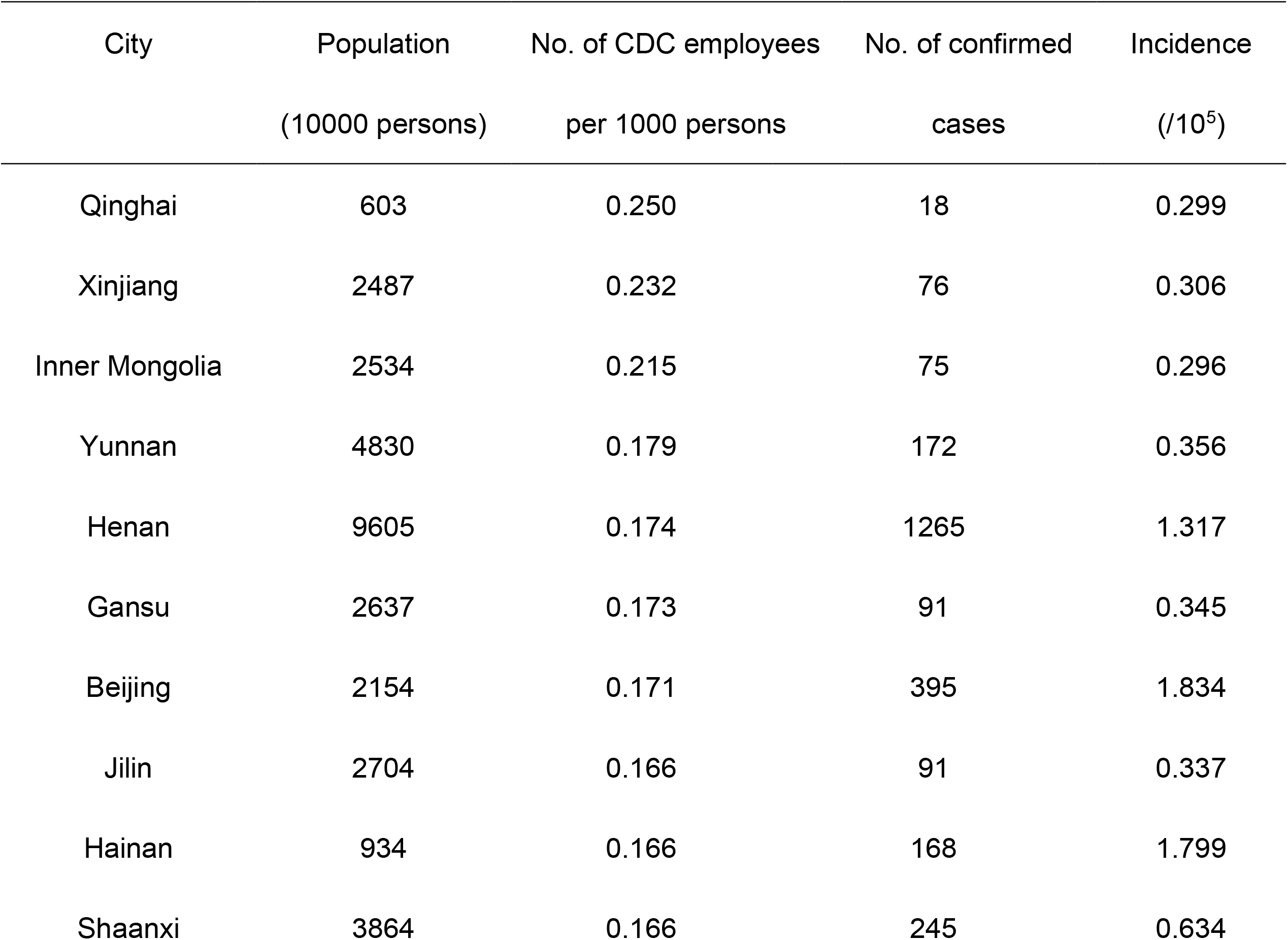
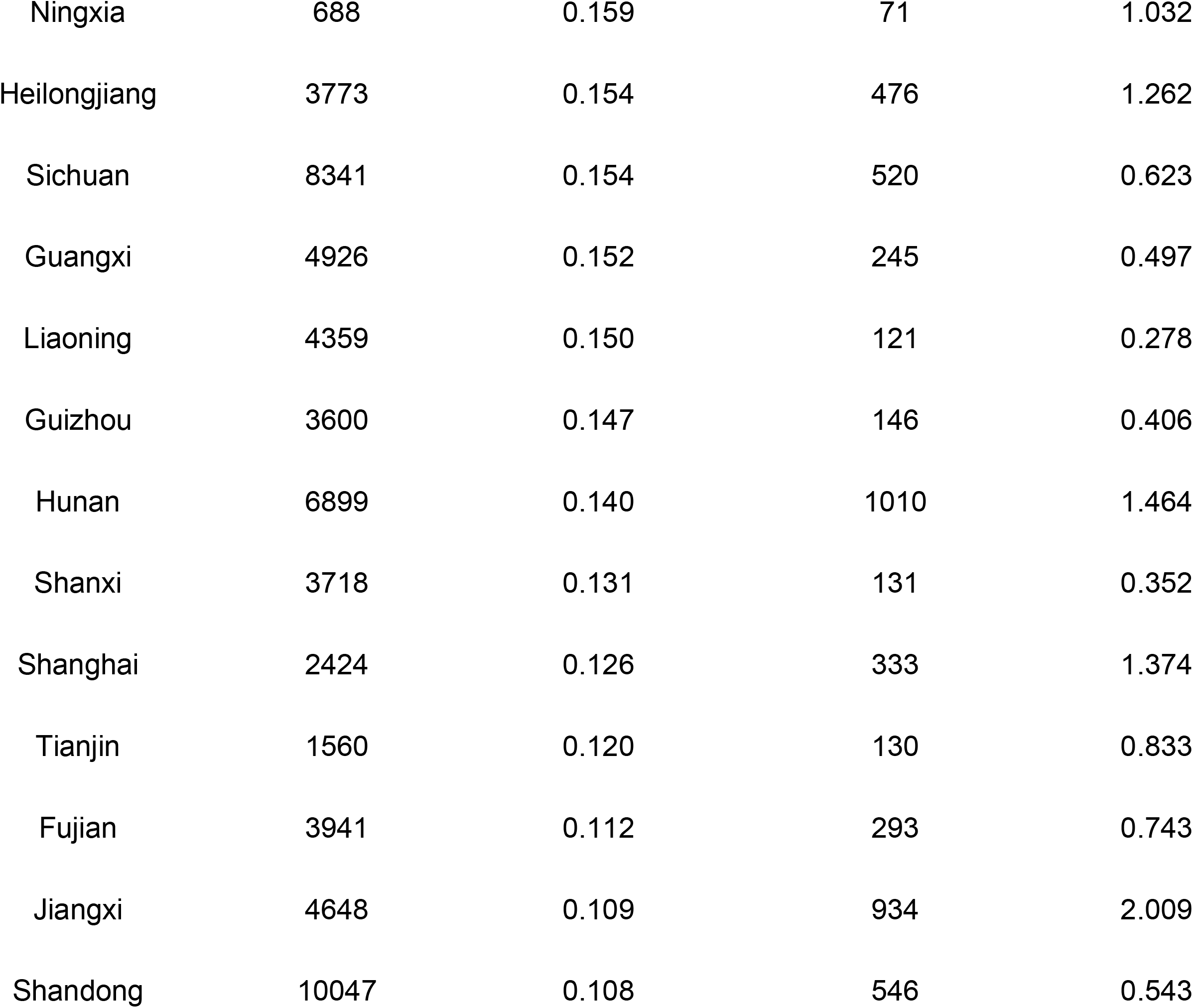
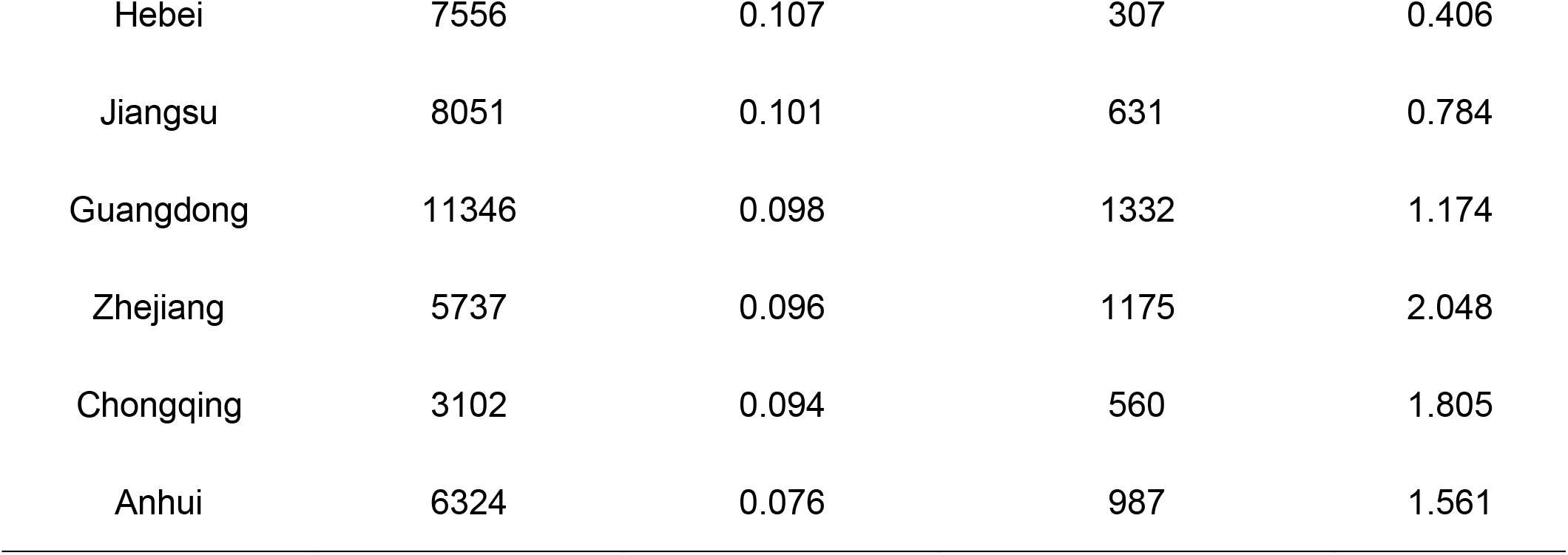
The Incidence of COVID-19 and CDC Employees per 1,000 Persons in Different Provinces

